# Advancing Sewage Sample Surveillance with OmiCrisp: A CRISPR-Based Platform for Variant-Specific SARS-CoV-2 Detection

**DOI:** 10.1101/2024.05.28.24307854

**Authors:** BS Kruthika, Bidipta Roy, Varsha Shridhar, Vaijayanti Gupta, Vijay Chandru, Reety Arora

## Abstract

The detection and differentiation of SARS-CoV-2 variants are vital for disease management and timely patient isolation. Although qRT-PCR has been the standard method, our OmiCrisp assay has shown superior results in identifying Omicron and non-Omicron variants. The World Health Organization advises ongoing sewage surveillance to monitor community viral load, accounting for both symptomatic and asymptomatic individuals. In a previous study, our CRISPR-based assay achieved a 99% concordance with qRT-PCR in sewage samples. We expanded this research by analyzing 182 sewage samples from Bangalore during the JN.1 (BA.2.86.1.1) variant outbreak. Using the OmiCrisp assay, 153 samples (84.06%) tested positive, 16 (8.79%) were negative, and 13 (7.14%) were low positive. Results indicated a significant reduction in positivity rates by the end of the study. The OmiCrisp assay thus proves to be an accurate and effective tool for community-wide surveillance of SARS-CoV-2 variants.

---

Since the outbreak of the SARS Cov2 pandemic, detection of the virus and its variants has been a crucial step in the management of the disease and timely isolation of the infected patients. Although qRT-PCR has been a go-to detection method from the beginning of the pandemic, our OmiCrisp assay has shown promising results in both the detection as well as in differentiating Omicron with the non-Omicron variants (Sharma S et al, 2023, Sharma S et al, 2024).

Apart from testing the symptomatic individuals, the World Health Organization also recommends a continuous surveillance of the sewage water. Waste water surveillance includes the virus shed by both symptomatic and asymptomatic individuals and thus gives an overview of the viral load in the community/population. In our previous study, we developed a CRISPR-based assay for the detection of the SARS Cov2 virus in both the blood samples of individuals as well as in the sewage water samples. We showed a 99% concordance of OmiCrisp with the qRT-PCR test in identifying SARS COV2 in sewage samples. The test also showed higher accuracy in identifying the Omicron variants as compared to the qRT-PCR test (Sharma S et al, 2023, Sharma S et al, 2024).

Owing to the results of our previous study, we have tested a further batch of 182 sewage samples collected between a period of Jan 2024 to February 2024. This was the time frame for the outbreak of the JN.1 (BA.2.86.1.1) variant in India. Since the JN.1 variant belongs to the Omicron lineage, we used the same set of guides (as used in our previous publication) to differentiate Omicron lineage versus non-Omicron in the current study.

Briefly, sewage water samples were collected and RNA isolation was done using NucleoDx RT RNA extraction kit (Cat No. NDX-RNA-021, Manufacturer: NeoDx Biotech Labs Pvt Ltd.) The extraction was performed at Molecular Solutions Care Health LLP, Bangalore. The samples were collected from open storm water drains from designated localities from across Bangalore city. The extracted RNA was stored at -20°C until use. For OmiCrisp assay, one step RT-PCR was performed with the commercially available TaqPath™ 1-Step Multiplex Master Mix (No ROX) (Catalog number - A28523, Applied Biosystems, USA) as per the protocol published previously (Sharma S et al, 2023, Sharma S et al, 2024). Trans-Cleavage assay was performed with Ascas12a enzyme (1μM), guide RNA (for ORF_Reference, ORF_Omicron, Ngene_Reference and Ngene_Omicron - 1μM each), fluorescence quencher-reporter (custom ssDNA_FQ reporter - /56-FAM/TT ATT /3IABkFQ/) procured from IDT(Integrated DNA Technologies, USA) and NEBuffer 2.0 (NEBuffer™ 2, New England BioLabs, USA). 5μl of RT-PCR product from each sample was added to the above mentioned reaction mix and incubated at 37°C in the Infinite 200 PRO multimode plate reader (Tecan Group Ltd., Switzerland) for 60 mins with fluorescence readings captured at every 2.5mins. The fluorescence readings at the 60 mins time-point were used for analysis. The data analysis and interpretation was done according to the previous publication (Sharma S et al, 2023, Sharma S et al, 2024).

A total of 182 wastewater samples were collected from 14 different localities. These samples were gathered over thirteen individual days spanning over a period of 51 days, from January 3rd, 2024, to February 23rd, 2024. The OmiCrisp assay was performed using the RNA isolated from each of the samples. The fluorescence readout for all the samples were plotted for ORF_ref, Ngene_ref and Ngene_om guides (Figure 2). All the samples with the read out higher than NTC+3SD were called out as positive. Of the 182 samples tested, 153 (84.06%) were positive, 16 (8.79%) were negative and 13 (7.14%) were low positive (read out marginally above the cut off value). There was a substantial reduction in the overall percentage positivity of the samples towards the end of the study period (Figure 3).

**Figure 1:**
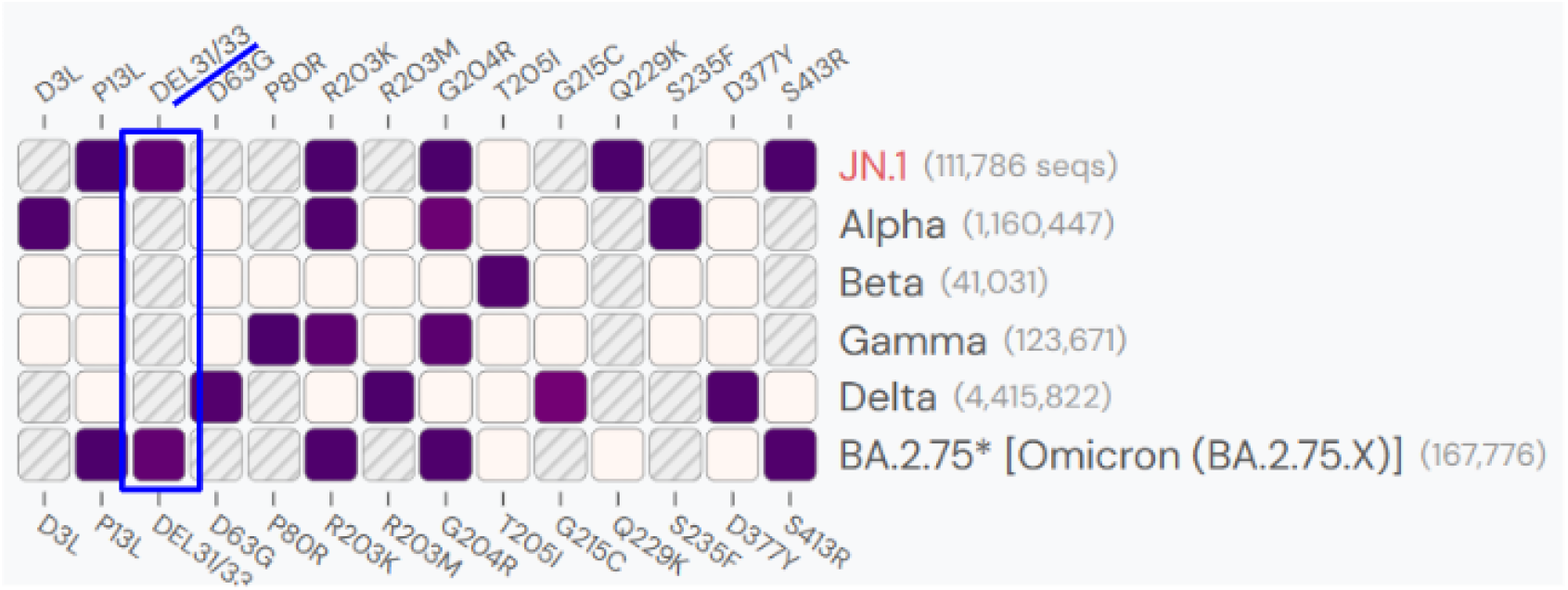
The heat map representing the mutations in Ngene across different variants of SARS-CoV2. The figure was generated on the website - outbreak.in. The figure shows the DEL31/33 mutation in Omicron and retained in the JN.1 variants had the same mutation in the Ngene.

**Figure 2:**
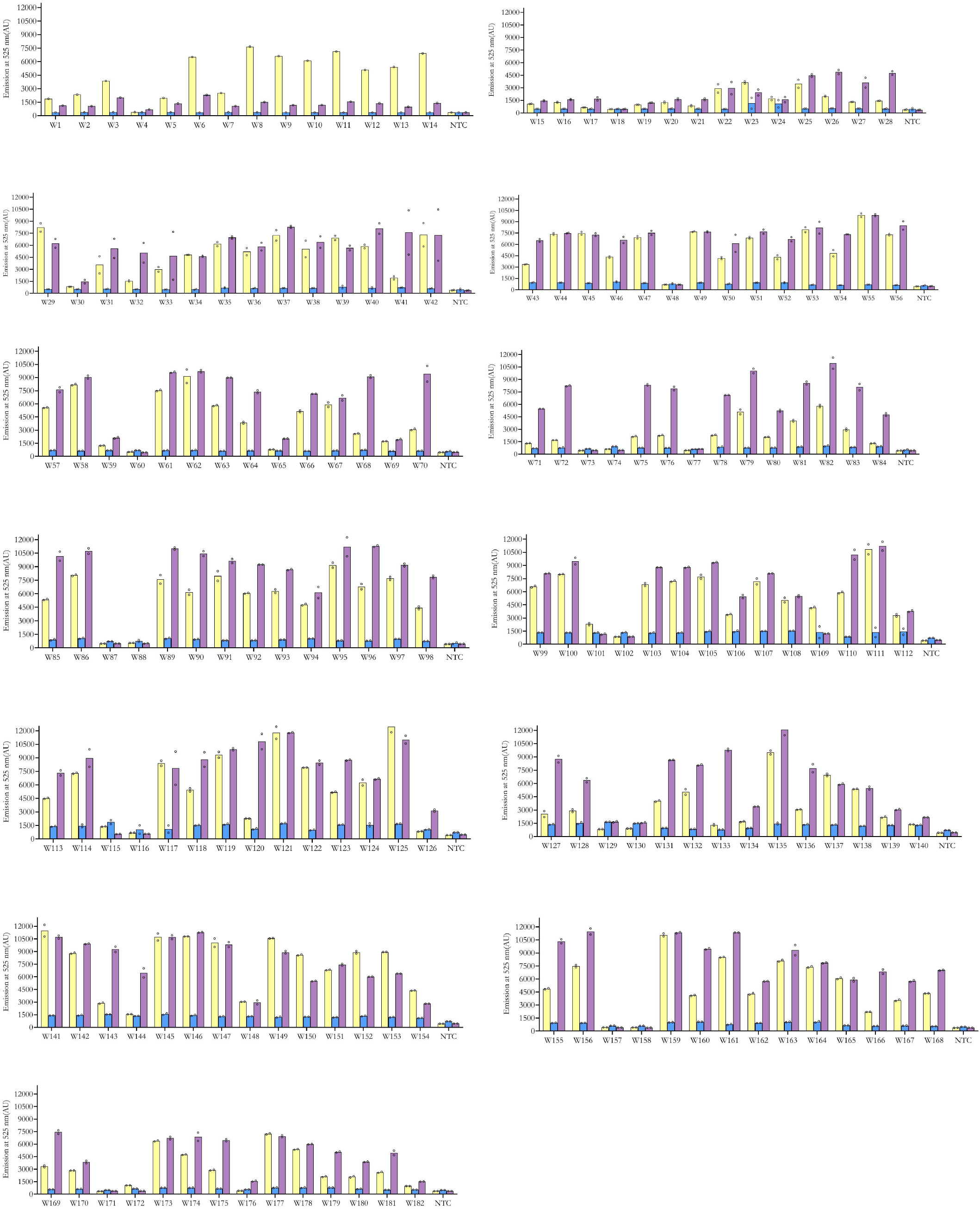
OmiCrisp assay testing for sewage water samples. The bar graph represents the fluorescence readout for the individual guide RNAs tested. The height of each bar in the graph represents the mean fluorescence intensity of the technical duplicates (except the first graph with sample number 1-14 - the testing was not done in duplicates). The circles that are on or above the bars represent the fluorescence intensity values of each of the technical duplicates.

**Figure 3:**
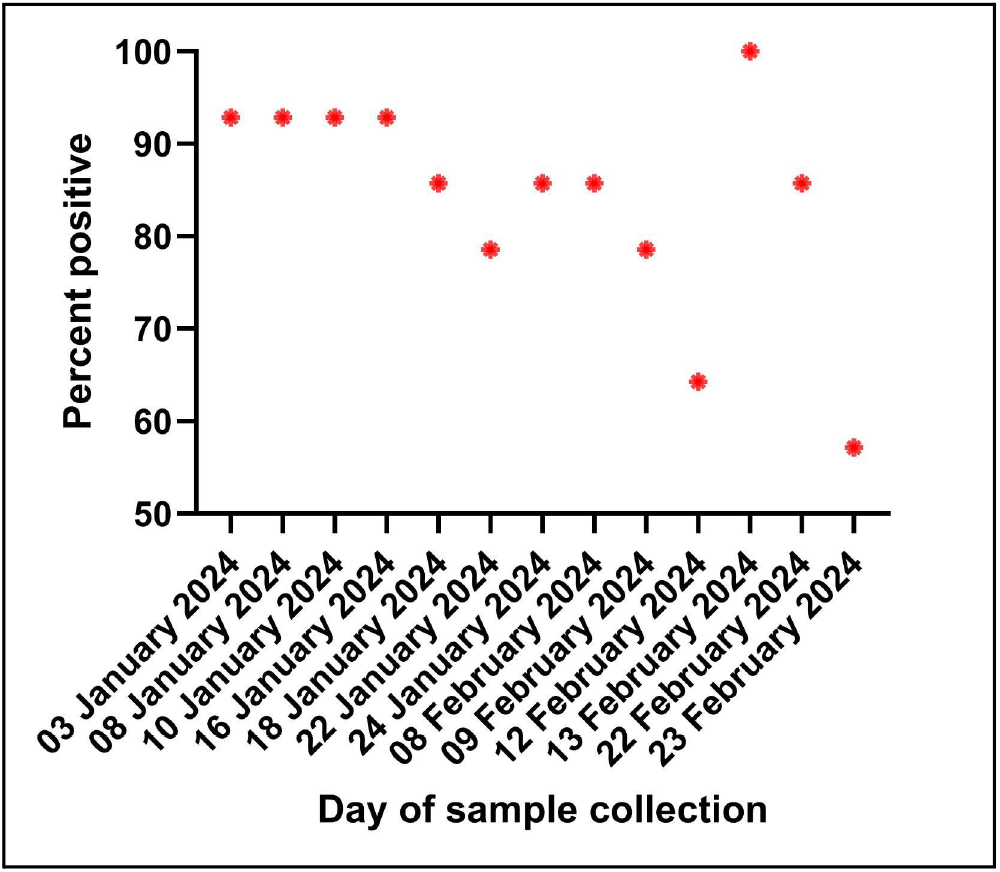
Graph representing the percentage positive on each day of sample collection. Each dot in the graph represents the percentage of samples showing positivity on each day of sample collection. A considerable decrease in the percentage positivity is observed towards the end of the study period.

We also observed that among the 14 localities examined, Jakkur and Thanisandra consistently tested negative for SARS-CoV-2 and the Omicron variant since the fourth and fifth day of sample collection, respectively. Further, the sample from day 10 showed low positivity followed by a significant positivity on the day 11. However, the sample from the consecutive week (day 12 and day 13) again tested negative for SARS-CoV-2 and the Omicron variant (Figure 4).

**Figure 4:**
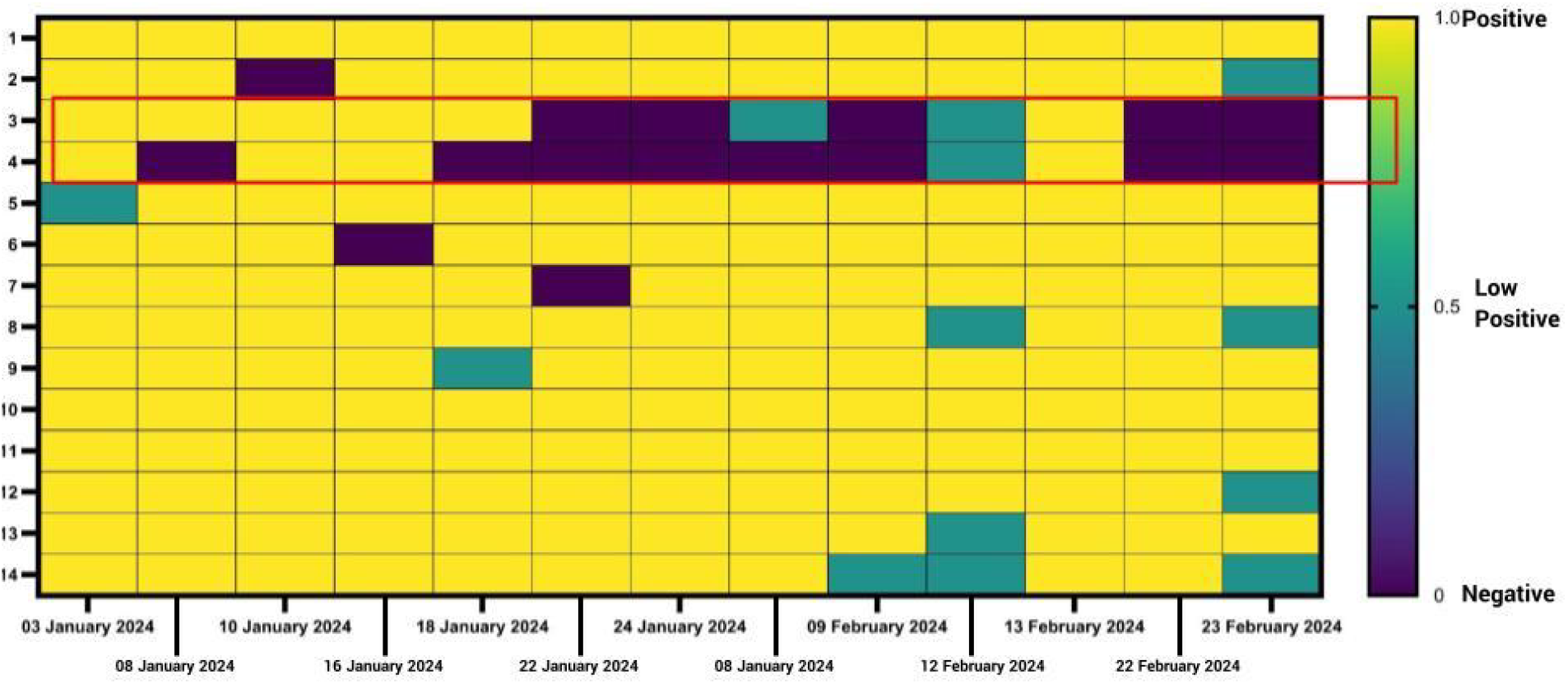
Heat map representing the positive, low positive and negative samples. The numbers on the Y-axis represent the localities (1-Vignana Nagar, 2-Banaswadi, 3-Jakkur, 4-Thanisandra, 5-Vishwanathanagenahalli, 6-HBR Layout, 7-Horamavu, 8-Bellandur, 9-Hongasandra, 10-Varthur, 11-BTM Layout, 12-Uttarahalli, 13-Jnanabharathi Ward, 14-Rajarajeshwari Nagar) and dates on the x-axis represent the date of the sample collection.

Overall, we show a successful application of our CRISPR Diagnostic assay OmiCrisp to accurately detect the presence of Omicron in waster-water collected across Bangalore. Therefore, the OmiCrisp assay proves to be a highly accurate and effective tool for the surveillance of SARS-CoV-2 variants at the community level.

## Data Availability

All data produced in the present work are contained in the manuscript

## Acknowledgements

We would like to express our gratitude to Block Chain for Impact to support this work and our continued efforts on waste water surveillance. We would also like to thank CDD (Consortium for DEWATS Dissemination India (CDD India) for help with collection of samples from open drains.

